# Aflatoxin M1 exposure and nutritional status of breastfeeding children 0-6 months in Makueni County, Kenya

**DOI:** 10.1101/2025.01.24.25321090

**Authors:** Isaac O. Ogallo, Dasel W. M. Kaindi, George O. Abong’, Alice M. Mwangi

## Abstract

Aflatoxin exposure in breastfeeding children has yet to be extensively explored in the southeastern region of Kenya, which is prone to aflatoxin outbreaks, and the assessment of potential risks may be underestimated. We determined the aflatoxin exposure of children 0-6 months and the outcome of weight-for-age z-scores as part of a larger cross-sectional study involving 170 randomly selected mother-child dyads. A semi-structured questionnaire was used to collect information on breastfeeding practices. Children’s weights were also measured. Breast milk samples of 48 lactating mothers and their children’s urine were collected from households whose food samples were collected for aflatoxin analysis. Aflatoxin levels were determined using enzyme-linked immunosorbent assays. The Statistical Package Software for Social Sciences (version 27) and Palisade’s @Risk software were used for analysis.

Exclusively breastfeeding children were 44.1% (75/170). The prevalence of aflatoxin M1 in breast milk was 77.1% (37/48) (concentration 35 ng/l; *SD*, 0.0), and the mean intake was 0.47 mg/kg b.w.t/day. All urine (100%) had aflatoxin M1 (mean 0.39 ng/ml (*SD*, 0.16). The exposure levels through breast milk were alarmingly high (margin of exposure < 10000) regardless of breastfeeding status, with complementary feeding potentially increasing the risks. Mothers’ diets significantly contributed to aflatoxin M1 in breast milk and urine (*p* = 0.01). Socioeconomic status, household size, and age of lactating mothers were not significant predictors of aflatoxin M1 exposure (*p* > 0.05). Aflatoxin M1 exposure did not influence the weight-for-age z-scores (*pall* > 0.05). The results indicate high exposure levels among children 0-6 months, and breastfeeding practices could be one of the under-evaluated risk factors.

## Introduction

Breastfeeding children between the ages of 0 and 6 months depend on breast milk as their primary nutrient and energy source for their growth and development. To date, this code of practice remains the norm among lactating mothers as breast milk is considered reliable, safe, and adequate during exclusive breastfeeding of children [1]. The insistence of this practice is because children under the age of six months have weak immune body systems and delicate digestive systems that cannot efficiently handle a diverse of foods, particularly solid foods [2]. However, in addition to breast milk, some children are also introduced to pre-lacteal feeds, including soft porridge, milk formula, and bovine milk. While breast milk is considered safe and more nutritious for children, the quality of breast milk depends significantly on mothers’ dietary intakes [1].

With this analogy, we posit that the consumption of foods susceptible to aflatoxin contamination by lactating mothers can potentially compromise the safety of breast milk and could put the health of breastfed children, particularly those below the age of 6 months, at risk due to the potency of aflatoxins even in smaller doses. A study [3] states that once aflatoxins are ingested, they are rapidly absorbed in the gut, degraded in the liver, transformed into residues, and later transferred to body fluids and tissues. Some are eliminated in the urine [4]. Though the transfer mechanism to body fluids is still unknown, the process involves enzymes in the liver that break down aflatoxins into toxic aflatoxins M1 and M2 [3], which can be detected in body fluids like milk, blood, and urine within 72 to 96 hours post-feeding [5]. A direct relationship between the amount of aflatoxin ingested and the amount of aflatoxin detected in dairy milk has been observed, with the levels shown to vary greatly depending on the concentrations of aflatoxin in the feeds, the amount of the feed consumed, the duration of the consumption, and the prevailing season [6].

Similarly, studies have determined children’s exposure to aflatoxin intake. Of interest to this study is exposure through breast milk to assess the safety of infants and young children in this aflatoxin-prone region. Past literature studies, including [7–12] conducted across age groups of children in different areas, reported varying levels of aflatoxin exposure in breast milk among lactating mothers. Nonetheless, all the studies agreed on the importance of determining toxicant levels in breast milk to control the transfer of chemicals to infants. Some of the recent studies on human breast milk include a study in Turkey [13], Serbia [14], Pakistan [15], Lebanon [16], Iran [17], and India [18]. Even though results were varying, they all pointed out the need to monitor levels of aflatoxin exposure in breastfeeding children.

A high concentration of aflatoxin has been associated with acute aflatoxicosis, while prolonged intake in small doses has been associated with chronic aflatoxicosis [19]. Acute aflatoxicosis is fatal and causes hemorrhage, edema, and acute liver damage. On the other hand, chronic aflatoxicosis is associated with the alteration of DNA, resulting in the occurrence of cancer, birth abnormalities in fetuses, malnutrition, and immune suppression in humans [3]. A negative correlation with anthropometric indicators (weight-for-age, height-for-age, and weight-for-height) on young children has been reported in some studies, including [20–22]. Results of Kang’ethe and colleagues [23] among breastfed children under five years showed aflatoxin exposure in urine and breastmilk was positively associated with higher malnutrition rates in Makueni County, Kenya. Similarly, a study [24] among children between 6 to 12 years in Makueni County, Kenya, also showed a negative association between aflatoxin B1-lysine and weight-for-age of children. Zhou and colleagues [25] concluded that aflatoxin interferes with the digestion and metabolism of several elements, including protein synthesis.

Besides previous studies [9, 23], there is a need to provide more information regarding aflatoxin exposure in children, particularly those below six months in Kenya who are recommended to be exclusively breastfed. Furthermore, most studies give little consideration to the role of prevailing breastfeeding practices, such as breastfeeding frequency, breast milk intake levels, and the influence of pre-lacteal feeds supplemented alongside breast milk for non-exclusively breastfeeding children on the aflatoxin exposure levels, particularly in this study region where high prevalences of aflatoxin have been reported. This could underestimate the assessment of potential risks to children under six months who rely on breast milk as the primary food source. Therefore, more research is needed to fully understand and mitigate the risks of aflatoxin exposure among children of this age group.

## Methods

### Sampling Procedure

The sampling procedure used in this study was adopted from Ogallo et al.[26]. The breastfeeding children 0-6 months were part of the 170 sampled lactating mother-child dyads whose sample size was determined using the Fisher et al. formula [27]. The number of breast milk and urine samples collected for aflatoxin M1 analysis (n = 48, each) was based on the 48 households where cooked food samples were picked for total aflatoxin and aflatoxin B1 analysis described in [26].

## Data Collection

### Breastfeeding practices of lactating mothers

A pretested semi-structured questionnaire was administered to collect data on breastfeeding practices. Lactating mothers were asked about the frequency with which they breastfeed their children during the day and night, the time they initiated breastmilk, whether their children were exclusively or non-exclusively breastfeeding, and the type of complementary foods they gave to their non-exclusively breastfeeding children.

Breast milk intake was also determined among breastfeeding children in the study. The maternal test weighing method, as described in [28] with slight modifications, was adopted to suit this study. This was conducted on lactating mothers whose food samples were picked for total and aflatoxin B1 analysis. Weight measurements for mothers before and after breastfeeding were taken using a two-decimal digital bathroom scale (arboleaf^®^, Japan). Loss in weight was taken to represent quantities of breast milk consumed by breastfeeding children, and measurements were made to the nearest 1g. A conversion factor of 1.03 g/ml (breast milk density) was used to express the recorded loss of weight (g) into volume (ml). To get the total quantity of breast milk consumed per day per breastfeeding child, the results were multiplied by the total frequency of breast milk feedings within a typical 24-hour period as reported by the lactating mothers. This is illustrated using the formula:

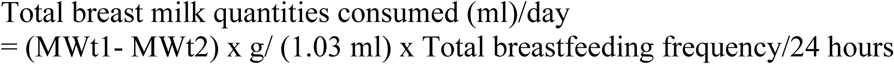

MWt1 is the maternal weight before breastfeeding, MWt2 is the maternal weight after breastfeeding, and 1.03g/ml is the density of breast milk. Breast milk intake per child was determined by dividing the total breast milk quantities consumed per day by the body weight of the breastfeeding child. The result was expressed in volume of breast milk (ml)/ kg b.w.t/day.

### Weight-for-age z scores of breastfeeding children

The weight of breastfeeding children was determined by first determining the weight of lactating mothers and getting the difference from the second weight of the same mothers measured standing on a scale while carrying the baby. A digital bathroom scale was used, and measurements were done to the nearest 0.01 kg. The age and weight of breastfeeding children were imported into WHO Anthro software (version 3.2.2). The weight-for-age z-score was compared against the WHO population standard age group below six months. Weight-for-age z-scores below -2 SD were considered underweight, while z-scores above -2 SD were considered normal in the study.

### Breast milk samples

Breast milk samples were collected from the lactating mothers a day after picking food samples. This was done with the help of female community health workers. Mothers first washed their hands, then cleaned and rinsed their breasts before expressing at least 10 ml of breast milk into a cryovial tube fitted with a Teflon cap. The breast milk was transferred into a cooler box and stored at about 4°C. Samples stayed for a maximum of two days before being taken to the laboratory. Storage was done in a deep freezer at -18°C before analysis. A total of 48 breast milk samples were collected for analysis.

### Urine samples

Instructions on how to collect early morning urine were given to lactating mothers a day (during breast milk) before the actual collection of urine samples. However, since it was challenging to collect mainstream urine from children 0 to 6 months, mothers were requested to collect urine from under wrappings (napkins or clothing) used on babies. Diapers were discouraged since they retain more urine than napkins and apparel. Lactating mothers were required, just after babies woke up in the morning (from 5 am onwards), to change the wet wrappings used at night, clean or dry the babies, wrap them again using a clean, dry napkin or clothing, and wait for the babies to pass urine. Once babies pass urine, mothers wrung the under wrappings, let urine drip into a plastic container provided (at least 10 ml), and transfer them to sterilized cryovials tubes. The collected urine samples were picked by the principal investigator and transferred into a cooler box at about 4°C for a day. Samples were taken to the laboratory the following day (six hours from the data collection site). Storage was done in a deep freezer at - 18°C before analysis. A total of 48 urine samples were collected for analysis.

## Analytical methods

### Determination of Aflatoxins M1 in the breast milk of lactating mothers

Aflatoxins M1 in breast milk was determined using Ridascreen^®^ Aflatoxin M1 ELISA kit (r-Biopharm, Darmstadt, Germany). A manual procedure was adopted for analysis. Breast milk samples (5ml) were centrifuged for degreasing and separation with upper-fat layers at 10 min/3500g at 10°C, and cream was removed by aspiration. Samples were diluted with 35% methanol (1:9) and put in the microwell. The antibody of 100 µL was added to the wells and incubated for 15 minutes. Approximately 100 µL of the diluted breast milk sample and standards were used per well and incubated for about 30 minutes at room temperature. The wells were washed using a 250 µL buffer solution. Conjugate of 100 µL was added, left to incubate for 15 minutes, and washed with 250 µL using phosphate buffer solution. Chromogen of 100 µL was added to the wells and left to incubate for 15 minutes. A stop solution of 100 µL was added to each well, and the reading was done within 15 minutes. Absorbance was determined at 450 nm. The lower detection limit was set at 5 ng/l.

### Determination of aflatoxin M1 intake in breast milk among breastfeeding children

Dietary aflatoxin intake was determined by multiplying the concentration of aflatoxin M1 in breast milk with estimated quantities of breast milk consumed in a day by breastfeeding children. The result was then divided by the body weight of breastfeeding children as shown in the formula:

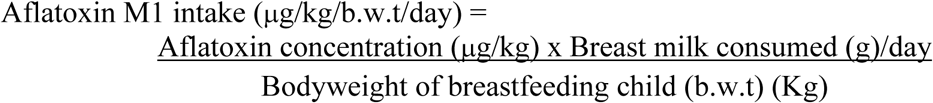

### Determination of margin of exposure of breastfeeding children to aflatoxin M1 intake

The margin of exposure was derived by taking the benchmark dose level (BMDL) of aflatoxin M1 and dividing it by the estimated aflatoxin M1 intake of breastfeeding children. A potency factor of 0.1 relative to Aflatoxin B1 BMDL_10_ of 0.17 μg/kg/b.w.t/day was applied for aflatoxin M1 in breast milk (European Food Safety Authority (EFSA) 2007). As a result, 0.017 μg/kg/b.w.t/day was used to assess the margin of exposure levels of breastfeeding children with a cut-off of greater than or less than 10000 MOE (EFSA, 2005) used to assess the risk levels of breastfeeding children using the formula:

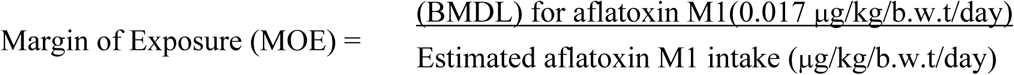

### Determination of Aflatoxin M1 in the urine of breastfeeding children

Aflatoxin M1 in urine was determined using the Aflatoxin M1 (urine) ELISA kit (Helica Biosystem Inc, California, USA). Urine (10 ml) was mixed with 40 ml of deionized water and filtered using a glass microfiber filter paper. Aliquots of urine standards and samples were further diluted with distilled water in the ratio of 1:20. Urine standards and samples (100 µL) were placed in microwells and buffered with 200 µL Phosphate Buffer Saline reconstituted with 0.05% Tween solution. After mixing, an antibody was added to each microwell and incubated for 1 hour at about 25°C. Tetramethylbenzidine stop solution was used on hose-radish-phosphate to stop the reaction. Reading was determined at 450 nm, and color was expected to change from blue to yellow. The detection limit was set at 0.15 ng/ml.

### Statistical Analyses

The statistical analysis methodology for this study is described in Ogallo et al. [26]. It was executed using Statistical Package Software for Social Sciences (SPSS version 27). Data mainly included breastfeeding practices, aflatoxin levels in breast milk, urine samples, and weight-for-age z-scores of breastfeeding children. Variables used in Ogallo et al. [26] on lactating mothers and assumed could influence breastfeeding children’s exposure were also utilized in this study. Data was first subjected to descriptive analysis. The difference between groups was determined using student t-test (*t*) for normally distributed data and Mann-Whitney *U* for non-normally distributed data. Statistical difference between more than three groups was determined using Analysis of variance (ANOVA) (*F*-test) for normally distributed data and the Kruskal-Wallis H-test for non-normally distributed data. Bonferroni’s Chi-square post hoc test was used for multiple pair comparisons of ranked variables, while Tukey’s b was used for the post hoc ANOVA test. Pearson (*r*), Kendall tau-b (*t_b_*), and Spearman (*rho*) were used to determine the correlation between normal continuous, non-normal continuous, and ranked variables, respectively.

In contrast, Chi-square (χ^2^ test) was used to determine the association between categorical variables. Simple and multiple linear regressions were used to determine significant predictors of aflatoxin concentration levels in analyzed foods, breastmilk of lactating mothers, urine of breastfeeding children, and outcome of weight-for-age z-scores. A significant level was set at p<0.05. @Risk software version 8.2 was used to determine the regression coefficient of each food on total aflatoxin and aflatoxin B1 intake in the study.

## Ethical consideration

This study complies with the ethical standards and procedures outlined in Ogallo et al. [26] as part of a more extensive study that evaluated Aflatoxin exposure of lactating mother-child pairs and the nutritional status of breastfeeding children 0-6 months in Makueni County. Ethical issues at all stages were considered in the study by obtaining ethical clearance (P454/08/2013) from Kenyatta National Hospital/the University of Nairobi-Ethical Review Committee (KNH/UoN-ERC). After approval of the research by the ethical committee, a meeting and discussion were held with the administrative and local community leaders, the Ministry of Health in charge, and community health workers to seek permission before conducting the study in the area. This was followed by subsequent awareness of the study at the village level, aiming to target lactating mothers and their spouses. A joint briefing was conducted where issues about the study safety, objectives, voluntary participation, and confidentiality of the study participants were highlighted before administering the study questionnaires. The survey was conducted between 7^th^ to 11^th^ of April 2014. Mothers willing to participate in the study signed written informed consent during data collection permitting the interviews and sample collection of food, breast milk, and their children’s urine and body weight measurements. However, mothers were also free to discontinue participating in the survey even after consent.

## Results

### Breastfeeding practices of lactating mothers in the study Breastfeeding practices of lactating mothers

The mean age of breastfeeding children (*n* = 170) was 3.8 months (*SD*, 1.5; range, 0.49-5.75 months). Less than half (44.1%) were exclusively breastfeeding. The average age for introducing other foods/or liquids alongside breast milk was 3.34 months (*SD*, 1.3). About half (48.4%) of non-exclusive breastfeeding mothers introduced complementary foods/liquids at the age of four months, 15.8% at the first month, 13.7% at the third month, and 10.5% for both the second and fifth months. Only 1.1% of the mothers introduced complementary feeds less than a month after delivery. All non-exclusively breastfeeding mothers (*n* = 95) mentioned animal milk as one of the most commonly used complementary foods, followed by 90.5% millet porridge, 87.4% maize porridge, 78.9% mixed flour porridge, 53.7% mashed *ugali*, and 46.3% sorghum porridge. However, maize porridge and animal milk (cow/goat) were the most frequently consumed complementary foods per week by 100% of non-exclusively breastfed children, followed by 73.7% stiff solid flour paste known as ‘*ugali.’* The least frequently consumed complementary food per week by non-exclusively breastfeeding children was millet porridge (11.5%), followed by sorghum porridge (16.8%) and mixed porridge (28.4%).

All breastfeeding children in the study were introduced to breast milk not more than 24 hours after delivery. Exclusively breastfeeding children, on average, consumed breast milk 17.2 times per 24-hour period, with a range of 9-26 times. On the other hand, non-exclusive breastfeeding children averagely consumed breast milk 13.9 times per 24-hour period, with a range of 7 to 25 times. The overall mean frequency over a typical 24-hour period was 15.3 times, with a statistical difference reported (Mann-Whitney *U*, *p* = 0.00).

About 73.3% and 68.4% of exclusively and non-exclusively breastfeeding mothers, respectively, desired to continue breastfeeding their children up to two years, while only 5.3% of exclusively and 4.2% of non-exclusive breastfeeding mothers desired to breastfeed their children up to six months. Those who desired to continue breastfeeding their children up to a year were 14.7% exclusively and 23.2% for non-exclusively breastfeeding mothers. A few exclusively (6.7%) and non-exclusively (4.2%) breastfeeding mothers desired to continue breastfeeding their children for up to three years.

Frequency consumption of complementary foods per typical day showed that 46.3% of the mothers fed their children maize porridge five times on a typical day, 23.2% thrice, 14.7% four times, and 15.8% twice. Animal milk was mostly consumed twice on a typical day by (49.5%) non-exclusively breastfed children., thrice by 28.4%, once by 14.7%, four times by 6.3%, and five times by 1.1%. On the other hand, 40.0% consumed mashed *ugali* twice on a typical day, 25.37% once, and 9.5% thrice. Sorghum porridge was mostly used once on a typical day (8.4%), followed by 4.2 thrice, 3.2% twice, and 1.1% four times. Millet porridge was consumed once per typical day by 6.3%, twice by 4.2%, and thrice by 3.2%. Mixed was consumed mostly twice by 13.7% of children, once by 6.3%, thrice by only 5.3%, and four times by 3.2%.

### Quantities of breast milk consumed per day by breastfed children in the study

The average quantity of breast milk consumed by breastfeeding children (*n* = 48) in the study was 543.3 ml/day (*SD*, 184.2; range, 233.0 to 922.3). Children aged between 0 to 1 month averagely consumed 583.6 (*SD*, 189) ml/day of breast milk, while those aged between >1 to 2 months averagely consumed 528.6 (*SD*, 203) ml/day. Children between >3 to 4 months averagely consumed 586.7 (*SD*, 207) ml/day, while those between >4 to 5 and >5 to 6 months consumed 491.3 (*SD*, 121) and 527.5 (*SD*, 176) ml/day, respectively (S1 Table). The quantities of breast milk intake, categorized by age groups and breastfeeding status, are presented in the S2 Table. Further analysis showed that exclusively breastfeeding children (*n* = 20) averagely consumed 588.4 (*SD*, 156.9) ml/day of breastmilk, while non-exclusively breastfeeding children (*n* = 28) averagely consumed 511.1 (*SD*, 197.9) ml/day. However, no significant difference was observed between them (*t*_46_ = 1.449, *p* = 0.154).

### Breast milk intake of breastfeeding children in Kibwezi West

The study’s mean breastmilk intake among breastfeeding children (*n* = 45) was 82.3 (*SD*, 31.7) ml/kg b.w.t/day, ranging from 31.6 to 157.8. The mean intake for exclusively breastfeeding children (*n* = 18) was 91.4 (*SD*, 34.1), while that of non-exclusively breastfeeding children (*n* =27) was 76.3 (*SD*, 28.9) ml/kg b.w.t/day. However, the means between the two breastfeeding groups were not significantly different (*t*_43_ =1.598, *p* = 0.117) (S3 Table).

### Weight-for-age z-score of breastfeeding children below six months in the study

The mean weight for children recruited in the study was 6.6 (*SD*, 1.9) kg, mode of 5 kg, and the range was between 2.3 and 11.0 kg. The mean weight for non-exclusively breastfeeding children (x̅ = 7.1, *SD* = 1.8 kg; *n =* 95) was significantly greater than the mean that of exclusively breastfeeding children (x̅ = 5.9, *SD* =1.8 kg, *n =* 75) at (*t*_168_ = -4.271, *p* = 0.00). Nutritional status based on weight-for-age z scores showed that 87.6% of children had normal weight (z-score >-2), while the rest 12.4% (13.3% for exclusively and 11.6% for non-exclusively breastfeeding children) were underweight (z-score <-2 SD). The mean weight-for-age z-scores for all breastfeeding children in the study was -0.1 (*SD*, 1.6), that of exclusively breastfeeding children was -0.4 (*SD*, 1.6; *n* = 75), while that of non-exclusive breastfeeding children was 0.1 (*SD*, 1.7; *n* = 95). A significant difference was observed between the two breastfeeding groups (*t*_168_ = -2.049, *p* = 0.042) (S4 Table ).

### Aflatoxin M1 in the breast milk of lactating mothers in the study

Aflatoxin M1 was detected in 77.1% (*n* = 48) breast milk samples of lactating mothers (90% for exclusively breastfeeding mothers [*n* = 20] and 67.9% for non-exclusively breastfeeding mothers [*n* = 28]). A prevalence ratio (PR) of 1.32 [95% C.I, 0.99 to 1.78] with an insignificant prevalence difference (Fisher’s exact sig. 2-sided, *p* = 0.09) was reported between the presence of aflatoxin M1 in breast milk exclusively and non-exclusively breastfeeding mothers in the study. However, of the positive samples (*n* = 37), slightly over three-fifths (61.8% [77.8% for exclusively (*n* = 18) and 52.6% for non-exclusively breastfeeding mothers (*n* = 19)]) were above 25 ng/l EU recommended limits set for infant milk. Overall mean concentration levels of aflatoxin M1 reported in the study, after correcting for normality using Shapiro-Wilk (*p* > 0.05), was 35 ng/l (*SD*, 0.0; range 5 to 77, *n =* 34), while that of exclusively and non-lactating mother were 38 ng/l (*SD* = 0.2; range 5 to 77, *n =* 16) and 32 ng/l (*SD* = 0.2; range 5 to 68, *n =* 18), respectively. No significant difference was reported between mean concentration levels of aflatoxin M1 in breast milk of the two breastfeeding groups of lactating mothers (*t*_32_ = 0.906, and *p* = 0.372).

### Aflatoxin M1 intake through breast milk among breastfeeding children in the study

Overall, the mean intake of aflatoxin M1 through breast milk was 0.47.μg/kg b.w.t/day (*SD*, 0.50; range, 0.0 to 1.7, *n* = 48). The highest mean intake was reported for children between >2 to 3 months (x̅ = 0.9 μg/kg b.w.t/day, *SD* = 0.57; range, 0.2 to 1.7, *n =* 6), while the lowest intake was reported for one child aged between >5 to 6 months (0.2 μg/kg b.w.t/day) (S5 Table).

Based on breastfeeding status, the mean aflatoxin M1 intake of exclusively breastfeeding children (*n =* 16) was 0.8 μg/kg b.w.t/day (*SD*, 0.50; range, 0.1 to 1.7). The highest intake (1.2 μg/kg b.w.t/day) in this group was reported for one child aged between 0 to 1 month, while the lowest (x̅ = 0.1 μg/kg b.w.t/day, *SD* = 0.09; range, 0.1 to 0.2) was reported for children aged between >3 to 4 months (*n* = 5) (S6 Table).

On the other hand, the overall mean aflatoxin M1 intake 0.6 μg/kg b.w.t/day (*SD*, 0.44; range, 0.0 to 1.7) was reported for non-exclusively breastfeeding children (*n* = 18). However, within this group, the highest mean aflatoxin M1 intake (x̅ = 0.8 μg/kg b.w.t/day, *SD* = 0.5; range 0.4 to 1.4) was reported for children aged between >3 to 4 months (*n* = 4), while the lowest intake (0.2 μg/kg b.w.t/day) was reported for a child aged between >5 to 6 months (S6 Table).

Generally, aflatoxin M1 intake between exclusively (*n* = 16) and non-exclusively breastfeeding children (*n* = 18) was not significantly different in the study (Mann-Whitney *U*, *p* = 0.198).

### Margin of exposure of breastfeeding children to aflatoxin M1 in breast milk

Aflatoxin M1 intakes (μg/kg/b.w.t/day) derived for positive breast milk samples showed that daily intakes of breastfeeding children (*n* = 34) were above 0.017 μg/kg/b.w.t/day benchmark level used in the study. Consequently, the margin of exposure (MOE) of all children regardless of breastfeeding status in the study, based on mean and 95^th^ percentile aflatoxin M1 intake through breastmilk, was lower than 10000 (European Food Safety Authority (EFSA) 2005) cut-off point (S7 Table).

### Aflatoxin M1 levels in the urine of children aged six months and below in the study

The overall mean of aflatoxin M1 in the urine of breastfeeding children (*n =* 48) was 0. 39 ng/ml (*SD*, 0.16), with a range of between 0.15 to 0.82 ng/ml. That of exclusively breastfeeding children (*n =* 20) was 0.35 ng/ml (*SD*, 0.13; range 0.15 to 0.61), while that of non-exclusively breastfeeding children was 0.42 ng/ml (*SD*, 0.18; range 0.18 to 0.82). Even though aflatoxin M1 in non-exclusively breastfeeding children’s urine was higher than in exclusively breastfeeding children’s, no significant difference was reported (*t*_46_ = -1.520, *p* = 0.135).

Within age groups of exclusively breastfeeding children (*n =* 20), the highest mean (0.43 ng/ml, *SD,* 0.16) and median (0.46 ng/ml) were reported for children aged between >4 to 5 months, while the lowest mean (0.16 ng/ml, *SD*, 0.01), and median (0.16 ng/ml) was reported for children aged between >1 to 2 months (S8 Table). On the other hand, within age groups of non-exclusively breastfeeding children (*n =* 28), the highest mean (0.54 ng/ml, *SD*, 0.19) and median (0.56 ng/ml) was reported for children aged between >1 to 2 months, while the lowest mean (0.22 ng/ml, *SD*, 0.49) and median (0.22 ng/ml) was reported for children aged between >2 to 3 months (S8 Table)

## Correlation of variables with aflatoxin occurrence in the study

### Correlation of variables with aflatoxin M1 levels in the breast milk of lactating mothers in Kibwezi West

A set of explanatory variables derived from Ogallo et al. (2023) study that this current study builds on was also correlated with the variables’ outcomes used herein. We assumed that they could also influence the variables’ outcomes of this study. They include socioeconomic status, dietary patterns, source of maize, maize storage, maize handling practices by the lactating mothers, and aflatoxin levels determined in foods consumed by the lactating mothers and whose breast milk, including the urine samples of their children, were collected.

A significant correlation was reported between total aflatoxin levels and aflatoxin M1 in breastmilk of lactating mothers in the study (*r* = 0.71, *p* = 0.00 for all mothers, *n =* 33; *t_b_* = 0.52, *p* = 0.005 for exclusively breastfeeding mothers, *n =* 16; and *t_b_* = 0.75, *p* = 0.00 for non-exclusively breastfeeding mothers, *n =* 17). Estimated intake of aflatoxin B1 statistically correlated with aflatoxin M1 in breastmilk of exclusively lactating mothers in the study (*t_b_* = - 0.56, *p* = 0.003) as opposed to that of non-exclusively breastfeeding mothers (*t_b_* = -0.07, *p* = 0.742). Estimates of total aflatoxin intake did not statistically correlate with the levels of aflatoxin M1 in breastmilk of lactating mothers in the study (*t_b_* = 0.10, *p* = 0.443 for all mothers, *n =* 33; *t_b_* = 0.75, *p* = 0.00 for exclusively breastfeeding mothers, *n =* 16: and *t_b_* = 0.27, *p* = 0.

139 for non-exclusively breastfeeding mothers, *n =* 17). In summary (Table 1), no socioeconomic and dietary consumption pattern variables were statistically correlated with aflatoxin M1 levels in the study’s breast milk of lactating mothers. Similarly, no statistically significant correlation was reported between variables that focused on where lactating mothers frequently sourced, where they stored, and how they handled maize before storage with levels of aflatoxin M1 found in breastmilk in the study.

**Table 1.** Correlation of variables with aflatoxin M1 levels in the breastmilk of lactating mothers in Kibwezi West.

| Variables | Aflatoxin M1 levels in breastmilk |  |  |  |  |  |
| --- | --- | --- | --- | --- | --- | --- |
|  | All mothers (*n* = 34) |  | EBF mothers (*n* = 16) |  | NEBF mothers (*n* = 18) |  |
|  | (*tb*) | p-value | (*tb*) | p-value | (*tb*) | p-value |
| **Sociodemographic** |  |  |  |  |  |  |
| Education level | 0.02<sup>*rho*</sup> | 0.893 | 0.32 | 0.225 | -0.16 | 0.518 |
| Age (mothers) | 0.04 | 0.721 | 0.01 | 0.964 | 0.04 | 0.818 |
| Household size | 0.03 | 0.818 | -0.04 | 0.853 | 0.25 | 0.182 |
| SES | -0.07 | 0.808 | -0.39 | 0.044 | 0.08 | 0.673 |
| **Dietary pattern** |  |  |  |  |  |  |
| Meals/day | -0.16 | 0.275 | -0.31 | 0.159 | -0.04 | 0.828 |
| Food score | -0.18<sup>*r*</sup> | 0.307 | 0.03 | 0.882 | 0.24 | 0.160 |
| WDD score | -0.03 | 0.80 | 0.04 | 0.851 | -0.19 | 0.323 |
| **Source of maize** |  |  |  |  |  |  |
| Market | -0.17 | 0.241 | -0.09 | 0.674 | -0.33 | 0.110 |
| Own cultivation | 0.10 | 0.508 | 0.07 | 0.745 | - | - |
| Other sources | 0.14 | 0.344 | 0.06 | 0.791 | 0.30 | 0.100 |
| **Storage practices** |  |  |  |  |  |  |
| Cleaning | 0.05 | 0.718 | 0.07 | 0.751 | -0.02 | 0.915 |
| Applying ash | -0.14 | 0.333 | - | - | -0.18 | 0.386 |
| Chemical | 0.03 | 0.827 | 0.21 | 0.341 | -0.04 | 0.859 |
| Drying | -0.03 | 0.608 | 0.07 | 0.745 | -0.11 | 0.574 |
| Nothing | -0.03 | 0.832 | -0.23 | 0.282 | 0.11 | 0.574 |
| **Place of maize storage** |  |  |  |  |  |  |
| Granary | -0.03 | 0.823 | -0.25 | 0.253 | 0.09 | 0.671 |
| Sacks | -0.02 | 0.815 | -0.10 | 0.634 | 0.18 | 0.390 |
| Buckets | -0.04 | 0.889 | 0.21 | 0.332 | -0.32 | 0.111 |
| **Aflatoxin** |  |  |  |  |  |  |
| AFB1 level <sup>a</sup> | -0.21<sup>*r*</sup> | 0.13 | -0.27 | 0.217 | -0.07 | 0.742 |
| AFT levels <sup>b</sup> | 0.71<sup>*r*</sup> | 0.000* | 0.52 | 0.005* | 0.75 | 0.000* |
| AFB1 intake <sup>a</sup> | -0.3 | 0.143 | -0.56 | 0.003* | -0.09 | 0.642 |
| AFT intake <sup>b</sup> | 0.10 | 0.443 | -0.10 | 0.587 | 0.27 | 0.139 |

Abbreviations: EBF, Exclusive breastfeeding; NEBF, Non-exclusive breastfeeding; SES; Socio-Economic status; AFB1, Aflatoxin B1; AFT, total aflatoxin; <sup>*ρ*</sup>, Spearman rho correlation; *t<sub>b</sub>*, Kendall's tau-b correlation; *r*, Pearson correlation.
<sup>a</sup> Sample sizes used after removing outliers for AFB1 (*n*= 26 for all mothers; *n*=12 for EBF; *n*=14 for NEBF).
<sup>b</sup> Sample sizes used after removing outliers for AFT (*n* = 33 for all mothers; *n*=16 for EBF mothers; *n*=17 for NEBF mothers).
\* Significant *p*<0.05

### Correlation of variables with aflatoxin M1 intake through breast milk of breastfeeding children in Kibwezi West

The age of lactating mothers did not statistically correlate with the intake of aflatoxin M1 through breastmilk among breastfeeding children in the study (*t_b_* = 0.01, *p* = 0.929). Likewise, education level (rho = 0.05, *p* = 0.687), household size (*t_b_* = 0.03, *p* = 0.794), socioeconomic status (*t_b_*= -0.13, *p* = 0.315), and age of breastfeeding children (*t_b_*= 0.004, *p* = 0.976) did not correlate with aflatoxin M1 intake in breast milk among breastfeeding children. Among dietary consumption variables, the consumption score of foods highly susceptible to aflatoxin contamination significantly and positively correlated with aflatoxin M1 intake of non-exclusively breastfeeding children (*t_b_* = 0.38, *p* = 0.031). On the other hand, women’s dietary diversity scores significantly and negatively correlated with intake of aflatoxin M1 among non-exclusively breastfeeding children (*tb =* -0.48, *p* = 0.010). Consumption scores of foods highly susceptible to aflatoxin contamination were also positively correlated with aflatoxin M1 intake among non-exclusively breastfeeding children (*tb =* 0.38, *p* = 0.031). No significant correlation was reported between aflatoxin M1 intake among breastfeeding children with total numbers of meals consumed per day by lactating mothers (*t_b_* = -0.007, *p* = 0.959) and time for initiating breastmilk with (*tb =* -0.10, *p* = 0.588). Total aflatoxin concentration levels in analyzed foods were significantly correlated with the intake of aflatoxin M1 in breast milk among breastfeeding children (*tb =* 0.31, *p* = 0.011).

Based on breastfeeding status, a statistically significant correlation was reported between total aflatoxin concentration levels and aflatoxin M1 intake of non-exclusively breastfeeding children in the study (*tb =* 0.38, *p* = 0.032) as opposed to non-exclusively breastfeeding children (*tb =* 0.02, *p* = 0.928). On the other hand, no significant intake was reported between aflatoxin B1 concentration levels with aflatoxin M1 intake among breastfeeding children (*t_b_* = -0.12, *p* = 0.378). However, the estimate of aflatoxin B1 intake of exclusively breastfeeding mothers significantly correlated with aflatoxin M1 intake of their breastfeeding children (*t_b_* = - 0.39, *p* = 0.037). This was not the case for non-exclusively breastfeeding mothers (*t_b_* = 0.16, *p* = 0.375). Conversely, the estimate of total aflatoxin intake of non-exclusively breastfeeding mothers significantly correlated with aflatoxin M1 intake of their breastfeeding children (*t_b_* = 0.49, *p* = 0.008) as opposed to exclusively breastfeeding mothers (*t_b_*= -0.14, *p* = 0.49). However, no statistically significant correlation was reported between the intake of aflatoxin M1 in breast milk and other variables in the study, as summarized in Table 2.

**Table 2.** Correlation of variables with aflatoxin M1 intake through breast milk among breastfeeding children in Kibwezi West.

|  | AFM1 intake (µg/kg/b.w.t/day) |  |  |  |  |  |
| --- | --- | --- | --- | --- | --- | --- |
|  | All (n=34) |  | EBF children (n=16) |  | NEBF children (n=18) |  |
| Variables | Correlation ($t_b$) | P-value | Correlation ($t_b$) | P-value | Correlation ($t_b$) | P-value |
| **Sociodemographic** |  |  |  |  |  |  |
| Age (mother) | 0.01 | 0.929 | -0.13 | 0.495 | -0.03 | 0.878 |
| Education level | 0.05<sup>rho</sup> | 0.687 | -0.15<sup>rho</sup> | 0.955 | 0.24<sup>rho</sup> | 0.339 |
| Household size | 0.03 | 0.794 | 0.18 | 0.354 | 0.08 | 0.656 |
| SES | -0.13 | 0.315 | -0.49 | 0.110 | 0.12 | 0.514 |
| Age (children) | 0.004 | 0.976 | -0.13 | 0.496 | 0.14 | 0.423 |
| **Dietary variables** |  |  |  |  |  |  |
| Meals/day | -0.007 | 0.959 | -0.21 | 0.329 | 0.22 | 0.276 |
| Time of initiating breastmilk | -0.10 | 0.588 | 0.15 | 0.445 | -0.21 | 0.253 |
| Food score | 0.09 | 0.466 | -0.14 | 0.458 | 0.38 | 0.031* |
| WDD score | -0.06 | 0.636 | 0.09 | 0.638 | -0.47 | 0.010* |
| **Source of maize** |  |  |  |  |  |  |
| Market | -0.14 | 0.334 | -0.25 | 0.248 | -0.09 | 0.657 |
| Shamba | 0.28 | 0.530 | 0.22 | 0.315 | 0.17 | 0.396 |
| Other sources | 0.07 | 0.642 | 0.10 | 0.634 | 0.09 | 0.657 |
| **Storage practices** |  |  |  |  |  |  |
| Cleaning | 0.13 | 0.366 | 0.10 | 0.634 | 0.24 | 0.243 |
| Applying ash | 0.02 | 0.878 | - | - | 0.14 | 0.500 |
| Apply chemical | 0.004 | 0.981 | 0.21 | 0.341 | -0.21 | 0.314 |
| Drying | 0.02 | 0.879 | 0.31 | 0.159 | -0.11 | 0.574 |
| No treatment | -0.08 | 0.570 | -0.38 | 0.079 | 0.00 | - |
| **Place for storing maize** |  |  |  |  |  |  |
| Granary | 0.003 | 0.983 | -0.10 | 0.638 | 0.17 | 0.396 |
| Sacks | -0.09 | 0.552 | -0.29 | 0.186 | 0.10 | 0.618 |
| Buckets | 0.06 | 0.685 | 0.26 | 0.225 | -0.22 | 0.288 |
| **Aflatoxin** |  |  |  |  |  |  |
| AFB1 level <sup>a</sup> | -0.12 | 0.378 | 0.03 | 0.891 | 0.00 | - |
| AFT level <sup>b</sup> | 0.31 | 0.011* | 0.02 | 0.928 | 0.38 | 0.032* |
| AFB1 intake <sup>a</sup> | -0.06 | 0.616 | -0.39 | 0.037* | 0.16 | 0.375 |
| AFT intake <sup>b</sup> | 0.17 | 0.183 | -0.14 | 0.469 | 0.49 | 0.008* |

Abbreviations: EBF, Exclusive breastfeeding; NEBF, Non-exclusive breastfeeding; SES; Socio-Economic status; AFB1, Aflatoxin B1; AFT, total aflatoxin; <sup>ρ</sup>, Spearman rho correlation; $t_b$ , Kendall's tau-b correlation; $r$ , Pearson correlation.
<sup>a</sup> Sample sizes used after removing outliers for AFB1 ( $n=26$ for all mothers; $n=12$ for EBF; $n=14$ for NEBF).
<sup>b</sup> Sample sizes used after removing outliers for AFT ( $n=33$ for all mothers; $n=16$ for EBF mothers; $n=17$ for NEBF mothers).
\* Significant $p < 0.05$

### Correlation of variables with aflatoxin M1 in the urine of breastfeeding children in Kibwezi West

No statistically significant correlation was reported between aflatoxin M1 in the urine of breastfeeding children (regardless of breastfeeding status) with age of lactating mothers (*r* = 0.18, *p* = 0.211), education level (*rho* = 0.00, *p* = 0.984), household size (*t_b_* = -0.01, *p* = 0.949), and socioeconomic status (*r* = -0.20, *p* = 0.173). No significant correlation was observed for these variables, even within exclusively and non-exclusively breastfeeding mothers. However, the age of exclusively breastfeeding children was positively and significantly correlated with the aflatoxin M1 in the urine of breastfeeding children (*t_b_* = 0.41, *p* = 0.017) as opposed to that of non-exclusively breastfeeding children (*t_b_* = 0.08, *p* = 0.953). Similarly, the socioeconomic status of lactating mothers was negatively correlated with aflatoxin M in the urine of their respective exclusively breastfeeding children (*t_b_* = -0.35, *p* = 0.041). However, no significant correlation was observed between dietary consumption variables of lactating mothers with aflatoxin M1 in the urine of breastfeeding children. Correlation with women’s dietary diversity score was *t_b_* = -0.06, *p* = 0.595, with consumption score of foods highly susceptible to aflatoxin contamination was *r* = -0.05, *p* = 0.761, and association with total number of meals per day was *t_b_* = 0.02, *p* = 0.854. Also, the age at which complementary foods were introduced was not statistically associated with aflatoxin M1 in the urine of non-exclusively breastfeeding children in the study (*t_b_* = 0.08, *p* = 0.588). A significant correlation was observed between the concentration of total aflatoxin in the study with aflatoxin M1 in the urine of breastfeeding children (*r* = 0.39, *p* = 0. 013).

Based on breastfeeding status, a significant correlation was noted between the total aflatoxin and aflatoxin M1 concentration in the urine of exclusively breastfeeding children in the study (*r* = 0.817, *p* = 0.00). Conversely, this was not the case for the non-exclusively breastfeeding children (*r* = 0.35, *p* = 0.115). No significant correlation was reported between aflatoxin B1 levels in the study with aflatoxin M1 in the urine of breastfeeding children (*r* = 0.27, *p* = 0.128). No statistical association was reported between aflatoxin intake in the study with aflatoxin M1 in the urine of breastfeeding children (total aflatoxin intake, *t_b_* = 0.13, *p* = 0.241; aflatoxin B1 intake, *t_b_* = -0.05, *p* = 0.673; and aflatoxin M1 intake, *t_b_* = 0.12, *p* = 0.320). No significant correlations were reported between aflatoxin M1 in urine with the maize source, maize storage practices, and place of storage variables, as summarized in Table 3.

**Table 3.** Correlation of variables with aflatoxin M1 in the urine of breastfeeding children in Kibwezi West.

|  | Aflatoxin M1 in urine |  |  |  |  |  |  |  |  |
| --- | --- | --- | --- | --- | --- | --- | --- | --- | --- |
|  | All children |  |  | EBF children |  |  | NEBF children |  |  |
| Variables | *n* | *t<sub>b</sub>* | *P-value* | *n* | *t<sub>b</sub>* | *P-value* | *n* | *t<sub>b</sub>* | *P-value* |
| **Sociodemographic** |  |  |  |  |  |  |  |  |  |
| Age (mothers) | 48 | 0.18<sup>r</sup> | 0.211 | 20 | 0.38 | 0.026 | 28 | -0.02 | 0.905 |
| Education level | 48 | 0.00<sup>rho</sup> | 0.984 | 20 | -0.25<sup>rho</sup> | 0.170 | 28 | 0.15<sup>rho</sup> | 0.288 |
| Household size | 48 | -0.01 | 0.949 | 20 | -0.18 | 0.297 | 28 | 0.09 | 0.562 |
| SES | 48 | -0.20<sup>r</sup> | 0.173 | 20 | -0.35 | 0.041* | 28 | -0.01 | 0.921 |
| Age (children) | 48 | 0.130 | 0.207 | 20 | 0.41 | 0.017* | 28 | 0.08 | 0.953 |
| **Dietary Pattern** |  |  |  |  |  |  |  |  |  |
| WDD score | 48 | -0.06 | 0.595 | 20 | 0.07 | 0.711 | 28 | -0.06 | 0.685 |
| Food score | 44 | -0.05<sup>r</sup> | 0.761 | 17 | 0.23 | 0.199 | 27 | -0.14 | 0.297 |
| Meals/day | 48 | 0.02 | 0.854 | 20 | -0.15 | 0.457 | 28 | 0.12 | 0.425 |
| Age complementary | - | - | - | - | - | - | 26 | 0.08 | 0.588 |
| **Aflatoxin** |  |  |  |  |  |  |  |  |  |
| AFT level | 41 | 0.39<sup>r</sup> | 0.013* | 19 | 0.82<sup>r</sup> | 0.00* | 22 | 0.35<sup>r</sup> | 0.115 |
| AFB1 level | 34 | 0.12<sup>r</sup> | 0.491 | 15 | -0.15<sup>r</sup> | 0.60 | 19 | 0.34<sup>r</sup> | 0.157 |
| AFM1 level (Breastmilk) | 34 | 0.27<sup>r</sup> | 0.128 | 16 | 0.49<sup>r</sup> | 0.055 | 18 | 0.19<sup>r</sup> | 0.457 |
| AFT Intake | 41 | 0.13 | 0.241 | 19 | 0.2 | 0.232 | 22 | 0.10 | 0.514 |
| AFB1 Intake | 41 | -0.05 | 0.673 | 19 | 0.15 | 0.523 | 22 | -0.12 | 0.455 |
| AFM1 intake | 34 | 0.12 | 0.320 | 16 | 0.20 | 0.278 | 18 | 0.98 | 0.570 |
| **Maize source** |  |  |  |  |  |  |  |  |  |
| Market | 48 | 0.02 | 0.876 | 20 | 0.10 | 0.620 | 28 | -0.02 | 0.889 |
| Shamba | 48 | -0.16 | 0.199 | 20 | -0.17 | 0.382 | 28 | -0.09 | 0.555 |
| Other sources | 48 | -0.03 | 0.811 | 20 | -0.05 | 0.816 | 28 | -0.02 | 0.908 |
| **Maize storage practices** |  |  |  |  |  |  |  |  |  |
| Cleaning | 48 | 0.04 | 0.715 | 20 | -0.04 | 0.85 | 28 | 0.09 | 0.589 |
| Applying ash | 48 | -0.01 | 0.963 | 20 | - | - | 28 | -0.05 | 0.758 |
| Chemical treatment | 48 | 0.1 | 0.422 | 20 | 0.30 | 0.129 | 28 | -0.04 | 0.810 |
| Drying | 48 | 0.01 | 0.966 | 20 | -0.12 | 0.129 | 28 | 0.04 | 0.789 |
| No treatment | 48 | -0.09 | 0.464 | 20 | -0.12 | 0.54 | 28 | 0.03 | 0.859 |
| **Place of storage** |  |  |  |  |  |  |  |  |  |
| Granary | 48 | -0.10 | 0.404 | 20 | -0.24 | 0.222 | 28 | -0.09 | 0.576 |
| Sacks | 48 | 0.12 | 0.311 | 20 | 0.01 | 0.970 | 28 | 0.24 | 0.143 |
| Buckets | 48 | 0.03 | 0.821 | 20 | 0.15 | 0.457 | 28 | -0.01 | 0.955 |

Abbreviations: EBF, Exclusive breastfeeding; NEBF, Non-exclusive breastfeeding; SES; Socio-economic status; WDD, women's dietary diversity; AFT, total aflatoxin; AFB1, Aflatoxin B1; AFM1, Aflatoxin M1; <sup>r</sup>, Spearman rho correlation; *t<sub>b</sub>*, Kendall's tau-b correlation; *r*, Pearson correlation.
\* Significant $p < 0.05$ .

### Correlation of variables and aflatoxin occurrence with weight-for-age outcome of breastfeeding children in Kibwezi West

The age of children was positive and significantly correlated with weight-for-age z-scores (*r* = 0.62, *p* = 0.00). Based on breastfeeding status, age was positively and significantly correlated with weight-for-age among non-exclusively breastfeeding mothers (*t_b_*= 0.56, *p* = 0.00) as opposed to their counterparts (*t_b_*= 0.28, *p* = 0.093). No significant correlation was observed between correlation of weight-for-age z-scores of breastfeeding children with age of lactating mothers (*r* = 0.13, *p* = 0.381), household size (*t_b_*= 0.04, *p* = 0.693), education level (*rho* = - 0.06, *p* = 0.67), and socioeconomic status (*r* = 0.03, *p* = 0.840) (Table 23). Similarly, total number of meals consumed by lactating mothers (*t_b_* = -0.06, *p* = 0706), the consumption score of foods highly susceptible to aflatoxin contamination (*t_b_* = 0.07, *p* = 0.649), and women’s dietary diversity score (*t_b_* = -0.12, *p* = 0.260) were not statistically correlated with weight-for-age of breastfeeding children in the study. No significant correlation was observed between breastfeeding practices and the study’s weight-for-age z-score of breastfeeding children. Correlation with time for initiating breastmilk was *t_b_* = 0.01, *p* = 0.940, age of introducing complementary foods was (*t_b_* = 0.22, *p* = 0.137), while the correlation with breastfeeding status of children was χ^2^ (1) = 0.09, *p* = 0.76. Correlations between weight-for-age z-score with aflatoxin B1 (*r* = 0.15, *p* = 0.405) and total aflatoxin levels (*t_b_*= 0.00, *p* = 0.993) in the study were not significant. Likewise, aflatoxin M1 in breastmilk of lactating mothers (*r* = 0.03, *p* = 0.887) and urine of breastfeeding children (*r* = 0.09, *p* = 0.350) were not statistically associated with weight-for-age z-scores in the study. Estimates of aflatoxin intake among lactating mothers did not correlate with the weight-for-age of breastfeeding children. A weak nonsignificant correlation was observed for aflatoxin B1 (*t_b_* = -0.07, *p* = 0.546), total aflatoxin intake (*t_b_* = -0.07, *p* = 0.509), and aflatoxin M1 intake through breastmilk (*t_b_* = 0.00, *p* = 0.796) (Table 4).

**Table 4.**
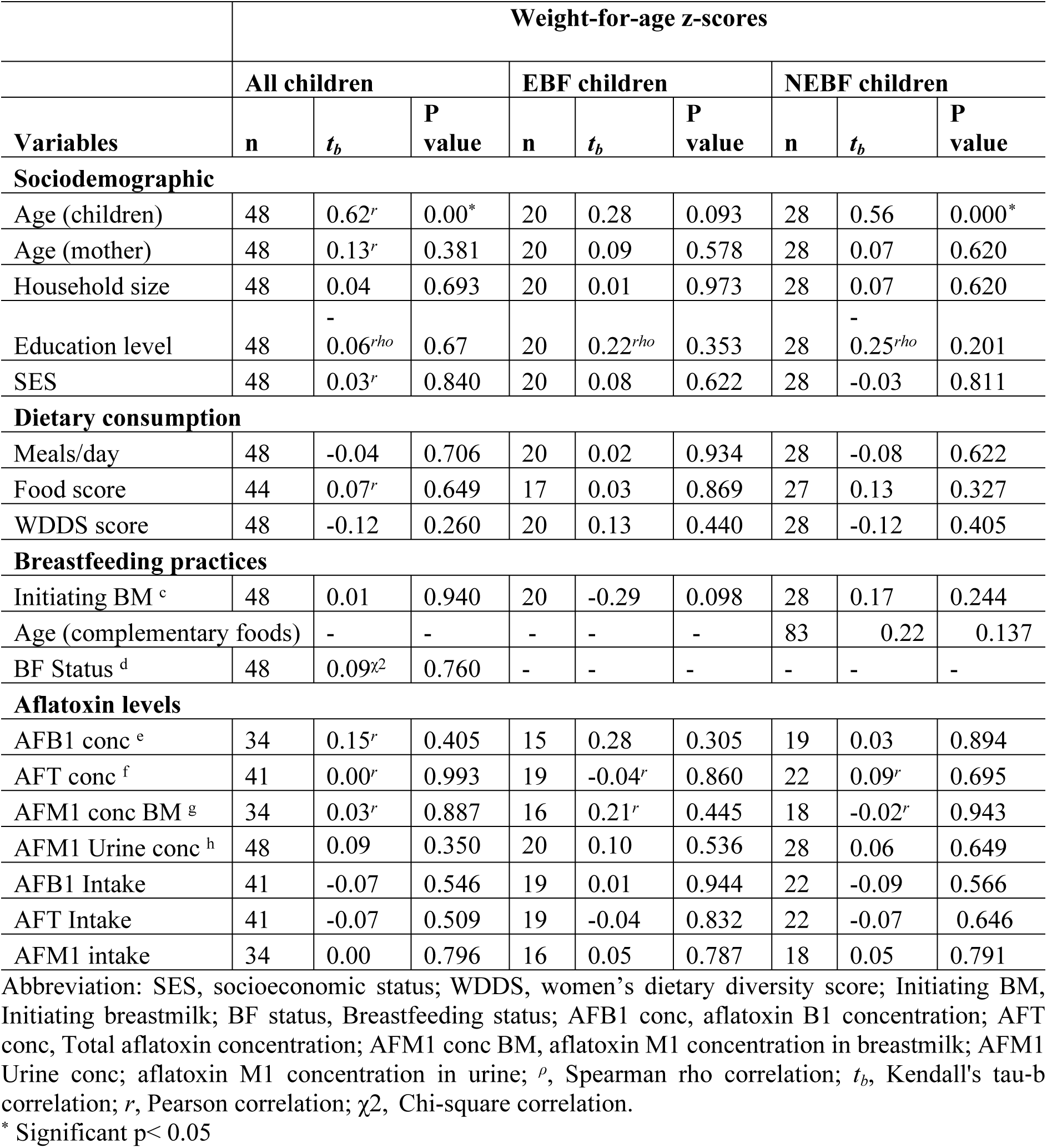
Correlation of variables with weight-for-age z-score of breastfeeding children in Kibwezi West.

### Predictors of aflatoxin and weight-for-age z-scores of breastfeeding children in Kibwezi West

Household size in a simple linear regression model (Adjusted R^2^=0.134, F_1,20_ = 4.250, and *p* = 0.052, enter method) was not a significant predictor of total aflatoxin in foods consumed by non-exclusively lactating mothers in the study (β= 2.06, *p* = 0.052). Also, women’s dietary diversity score (β= -0.27, *p* = 0.22) and sorting out of maize before storage (β= 0.36, *p* = 0.11) were not significant predictors of concentration of total aflatoxin in the study despite generating a significant predictor model (Adjusted R^2^ = 0.214, F_2,19_ = 3.853, *p* = 0.039) (Table 5). Similarly, socioeconomic status did not significantly contribute to aflatoxin B1 intake among lactating mothers in the study (β = 0.296, *p* = 0.06). However, the level of education significantly and negatively influenced estimates of aflatoxin B1 intake among exclusively lactating mothers in the study (β = -0.56, *p* = 0.01). On the other hand, women’s dietary diversity scores negatively influenced estimates of aflatoxin B1 intake among non-exclusively lactating mothers (β = -0.43, *p* = 0.04). Moreover, aflatoxin concentration in analyzed foods significantly contributed to levels of aflatoxin M1 in breastmilk of lactating mothers (β = 0.71, *p* = 0.00), while based on breastfeeding status, estimates of aflatoxin B1 intake in the study significantly influenced the levels of aflatoxin M1 in breastmilk of exclusively lactating mothers (β = -0.63, *p* = 0.01).

**Table 5.** Predictors of aflatoxin occurrence and weight-for-age z-scores of breastfeeding children in Kibwezi West.

|  | Unstandardized Coefficients |  | Standardized Coefficients |  |  |
| --- | --- | --- | --- | --- | --- |
| Predictors | B | Std. Error | Beta | t | Sig. |
| **AFT conc (NEBF mothers)** |  |  |  |  |  |
| Household size | 27.68 | 13.43 | 0.42 | 2.06 | 0.052 |
| **AFT intake (NEBF mothers)** |  |  |  |  |  |
| Women Dietary Diversity score | -1.35 | 1.05 | -0.27 | -1.28 | 0.22 |
| Cleaning maize before storage | 6.55 | 3.85 | 0.36 | 1.70 | 0.11 |
| **AFB1 intake (All mothers)** |  |  |  |  |  |
| Socioeconomic Status | 0.06 | 0.031 | 0.296 | 1.938 | 0.06 |
| **AFB1 intake (EBF mothers)** |  |  |  |  |  |
| Mother Level of Education | -0.23 | 0.08 | -0.56 | -2.81 | 0.01* |
| **AFB1 intake (NEBF mothers)** |  |  |  |  |  |
| Household size | -0.25 | 0.13 | -0.38 | -1.93 | 0.69 |
| Women Dietary Diversity score | -0.22 | 0.10 | -0.43 | -2.21 | 0.04* |
| **AFM1 conc BM (all mothers)** |  |  |  |  |  |
| AFT concentration | 0.00 | 0.00 | 0.71 | 5.64 | 0.00* |
| **AFM1 conc BM (EBF mothers)** |  |  |  |  |  |
| AFB1 Intake | -0.02 | 0.01 | -0.63 | -4.09 | 0.01* |
| AFT concentration | 0.00 | 0.00 | 0.49 | 3.18 | 0.07 |
| **AFM1 intake (All children)** |  |  |  |  |  |
| AFT concentration | 0.00 | 0.00 | 0.49 | 3.16 | 0.00* |
| **AFM1 intake (EBF children)** |  |  |  |  |  |
| AFB1 Intake | -0.38 | 0.19 | -0.47 | -2.00 | 0.07 |
| **AFM1 intake (NEBF children)** |  |  |  |  |  |
| Women dietary diversity | -0.18 | 0.08 | -0.47 | -2.31 | 0.04* |
| Food score | 0.13 | 0.07 | 0.36 | 1.78 | 0.10 |
| **AFM1 urine (all children)** |  |  |  |  |  |
| AFT concentration | 0.00 | 0.00 | 0.39 | 2.61 | 0.01* |
| **AFM1 urine (EBF children)** |  |  |  |  |  |
| Age (children) | 0.04 | 0.02 | 0.26 | 1.89 | 0.08 |
| Socioeconomic status | -0.01 | 0.06 | -0.16 | -1.14 | 0.27 |
| AFT concentration | 0.00 | 0.00 | 0.70 | 4.91 | 0.00* |
| **AFM1 urine (NEBF children)** |  |  |  |  |  |
| AFT concentration | 0.00 | 0.00 | 0.46 | 2.68 | 0.02* |
| AFT Intake | 0.02 | 0.01 | 0.25 | 1.34 | 0.20 |
| Women Dietary Diversity score | -0.12 | 0.06 | -0.32 | -1.97 | 0.72 |
| Food score | 0.01 | 0.01 | 0.18 | 1.25 | 0.24 |
| **WFA z-scores (all children)** |  |  |  |  |  |
| Age (children) | 0.78 | 0.14 | 0.62 | 5.41 | 0.00* |
| **WFA z-scores (EBF children)** |  |  |  |  |  |
| Age (NEBF children) | 0.88 | 0.16 | 0.73 | 5.50 | 0.00* |

Abbreviation: AFT, total aflatoxin; AFB1, Aflatoxin B1; AFM1, Aflatoxin M1; EBF, exclusively breastfeeding; NEBF, Non-exclusively breastfeeding; WFA; weight-for-age z-scores
\* Significant at $p < 0.05$

The study also found that total aflatoxin levels were a significant predictor of aflatoxin M1 estimate intake through breastmilk among breastfeeding children (β = 0.49, *p* = 0.00). The study also showed that the regression model containing women’s dietary diversity and consumption score of foods that are highly susceptible to aflatoxin contamination regarding aflatoxin M1 intake was significant (Adjusted *R^2^*= 0.023, *F* _(2,15)_ = 4.912, *p* = 0.023). However, women’s dietary diversity score was a significant predictor of aflatoxin M1 intake among non-exclusively breastfeeding children (β = -0.47, *p* = 0.04) as opposed to the consumption score of foods that are highly susceptible to aflatoxin contamination in the model (β = 0.36, *p* = 0.10). Furthermore, the total aflatoxin level was the major contributor of aflatoxin M1 in the urine of breastfeeding children in the study (β = 0.39, *p* = 0.01). Based on breastfeeding, regression model (Adjusted R^2^ = 0.698, F_3,15_ = 14.872, *p* = 0.000) showed that total aflatoxin (β = 0.70, *p* = 0.00) was a significant predictor of aflatoxin M1 in urine of exclusively breastfeeding children, as opposed to age of children (β = 0.26, *p* = 0.08) and socioeconomic status of mothers (β = -0.16, *p* = 0.27). Finally, age was the only significant predictor of weight-for-age z-score outcome among breastfeeding children in the study (β = 0.39, *p* = 0.01 for all children, and β = 0.73, *p* = 0.00 for exclusively breastfeeding children). The rest are summarized in Table 5.

## Discussion

### Breastfeeding practices of lactating mothers in Kibwezi West

Overall, this study’s exclusive breastfeeding rate (44.1%) was lower than the national rate of 59.9% [29]. This was unsurprising as Makueni County falls under Arid and Semi-Arid Land areas (ASALs) in Kenya, which are presumed to face several health and nutrition constraints that could directly or indirectly influence adherence to exclusive breastfeeding compared to non-ASAL areas. Indicative of these assertions are studies by Mohammed et al. [30] in Wajir County and Talbert et al. [31] in Kilifi County, classified as ASAL areas, also reported similar exclusive breastfeeding rates of 44.5% and 45%, respectively. However, the exclusive breastfeeding rate reported in this study was higher than those reported in similar aflatoxin exposure studies. A rate of 36.7% (combined rate) was reported in Tanzania [20], 28% in northern India [18], 35.4% in Nigeria [32] 40% in Lebanon [16]. The disparities among these rates could be due to varying degrees of constraints that lactating mothers face in exclusively breastfeeding their children. For instance, in this study, education status, occupational status, and age of breastfeeding children were all associated with the breastfeeding status of lactating mothers. These observations were similar to those of the Kenya Demographic Health Survey, 2014 [33] and Jamaa et al. [34].

All breastfeeding children in the study were introduced to breastmilk not more than 24 hours after delivery. This was higher than the 58% reported in Wajir County [34]. The rate was, however, higher but comparable to the national rate (92.1%) [33]. The average age for introducing other foods alongside breastmilk was 3.34 months. This was similar to those reported in Kilifi County (3 months) [31] but slightly higher than the 2.7 months reported in the western region of Kenya [35]. Complementary foods consumed in this study area (maize porridge, animal milk, and mashed *ugali*) were similar to those mentioned in other studies. In Kilifi County, maize porridge was the most frequently used complementary food [31]. In Makueni County, maize, sorghum, millet, and animal milk were mentioned by Kang’ethe et al. [23]. Wajir County widely used animal milk [34]. It is, however, surprising that over 60% of lactating mothers expressed the desire to continue breastfeeding for up to 2 years, almost matching the rate (60%) reported by KDHS (2014). Generally, a high non-exclusive breastfeeding rate characterized by frequent consumption of complementary foods in this study was seen as an additional risk of aflatoxin exposure among breastfeeding children, which agrees with the findings of Kang’ethe et al. [23]. The findings of this study further concur with those of Mehta et al. [18] and Ezekiel et al.[32]. These studies also concluded that non-exclusively breastfeeding children are exposed to higher levels of aflatoxin intake than those exclusively breastfed. However, we posit that besides determining the complementary foods frequently fed to the non-exclusively breastfed children, determining their dietary patterns, the aflatoxin levels in the foods they consumed, and in combination with aflatoxin in breast milk, could potentially give more accurate information regarding their level of exposure to aflatoxin intakes.

The average consumption quantity of breast milk in this study (559.6 g/day) was within the ranges of 525.3 to 793.1g/day reported in Tanzania [20]. The level was, however, below the reference consumption level of 750 g/day adopted in most aflatoxin M1 breast milk studies. A consumption quantity of 606.1 g/day was reported for exclusively breastfeeding children. They were lower than 841.2 g/day reported in western Kenya [36] and 750 g/day reported in northern India [18]. Despite non-exclusively breastfeeding children consuming other foods, it was surprising to report an insignificant difference between breast milk intake of exclusively and non-exclusively breastfeeding children. This shows that all breastfeeding children aged six months and below, regardless of breastfeeding status, are equally predisposed to aflatoxin M1 through breast milk in the study area. However, the statistical difference reported in breast milk intake between children age groups was expected. This intimates that age could be an underlying factor in determining the extent of exposure of breastfeeding children to aflatoxin M1 in breast milk.

### Aflatoxin M1 in breastmilk of lactating mothers in Kibwezi West

To the best of my knowledge, research on the occurrence of aflatoxin M1 in the breast milk of lactating mothers in Kenya is scanty. It was only conducted by Kang’ethe et al. [23] on mothers with children under 5 years old and Maxwell et al. [9] on pregnant mothers in Kenya. This study will be the third and the first to determine the presence of aflatoxin M1 in mothers who exclusively and non-exclusively breastfeed children aged six months and below in Kenya.

The prevalence of aflatoxin M1 in breast milk reported in this study (77.1%) was lower than 86.7% in Makueni but higher than 56.7% in Nandi [23]. The rates, however, were higher than the 28% reported by Maxwell et al. [9]. These rates confirm that the prevalence of aflatoxin in breast milk is as high as those reported in food samples in the study area. Comparison with 41% in northern India [18], 42% in Iran [37], 90% in Columbia [38], 93.8% in Lebanon [16], and 100% in Iran [17] show that the occurrence of aflatoxin M1 in the breast milk of lactating mothers is a widespread problem and vary from region to region. For exclusively breastfeeding mothers, 80% was reported in this study, against 18% in Nigeria [39] and 22% in Iran [21]. These high rates, especially for exclusively breastfeeding children, are alarming in the study area.

Based on the concentration of aflatoxin M1 in breast milk, 35 ng/L reported in this study was considerably higher than 8.46 ng/L and 0.02 ng/L reported in Makueni and Nandi, respectively, using ELISA [23]. However, ranges reported by Maxwell et al. [9] (5-1379 ng/L) in 121 breast milk samples were higher than the overall ranges reported in this study (5-78 ng/L). This outcome points out that aflatoxin occurrence in the breast milk of lactating mothers is an under-evaluated risk in Kenya. While it is considered that breast milk is the safest food for children below six months of age, this might not be the case in the study area. However, this problem also exists in other countries. The results of Mehta et al. [18] (3.9-1200ng/L, median 13.7 ng/L) were higher than the overall ranges reported in this study (5-78 ng/L). Pooled mean for Iran (5.85 ng/L) [37], and 4.31 ng/L for Lebanon [16] was lower than the mean of this study. The mean of aflatoxin M1 in the breast milk of exclusively breastfeeding mothers in this study (38.0 ng/L) was almost 40 times greater than the 2.0 ng/L levels reported in Ogun estate, Nigeria [32]. It was nearly six times greater than the 6.96 ng/L levels reported in rural areas of Iran [21]. However, mean levels reported in Tanzania (70 and 80 ng/L) during the rainy and dry seasons, respectively, by Magoha et al. [20] and 45 ng/L in Ecuador by Ortiz et al. [40] were higher than the values reported in this study. However, this study’s results were within the mean levels compiled by Fakhri et al. [41] for Africa. These comparisons with a high proportion (61.8%) of breast milk with aflatoxin M1 above 25 ng/L EU limits cause food safety and health concerns in the study area. Because of this, this study agrees with Kang’ethe et al. [23], who concluded that infant children in Makueni are also exposed to aflatoxin M1 through breast milk. This study confirms that breast milk in the study area is contaminated with aflatoxin. However, it adds that both exclusively and non-exclusively breastfeeding children aged six months and below in Makueni are equally exposed to aflatoxin M1 through breast milk.

### Intake of aflatoxin M1 in breastmilk among breastfeeding children in Kibwezi West

Intake of aflatoxin M1 in this study was generally high among children regardless of their breastfeeding status. The overall mean intake (0.47 μg/kg b.w.t/day) was higher than 0.006 and 1× 10^-6^ μg/kg b.w.t/day in Makueni and Nandi, respectively [23]. They were also higher than those estimated using dairy milk in Nairobi (0.004 μg/kg b.w.t/day) [22]. The result of this study was comparable on an age group basis with those of Hernández et al. [42]. However, the levels were higher than 0.012 μg/kg b.w.t/day reported in Tanzania [20], 0.069 μg/kg b.w.t/day reported in Lebanon [16], 0.003 μg/kg b.w.t/day reported in Serbia [14], and 3.04×10^-4^ μg/kg b.w.t/day reported in India [18].

The results of this study were consistently higher, thus pointing to the existence of high aflatoxin exposure among breastfeeding children aged six months and below in the study area. Consequently, a low margin of exposure value (<10000) showed that both exclusively and non-exclusively breastfeeding children in the study area were highly exposed to high levels of aflatoxin intake. These results confirm the findings of Coppa et al. [43], who reported a margin of exposure of 0.27 for African countries. Because of this, this study acknowledges that the margin of exposure of breastfeeding children to aflatoxin is high in African countries. In Kenya, this study is the first to estimate the margin of exposure of breastfeeding children to aflatoxin M1 through breast milk. However, margin exposure reported in Serbia [44] was greater than 10,000 for children aged between 1 to 9 years. Similarly, those reported in Italy for toddlers were below 10,000 but considerably higher than those reported in this study [45]. Because of this, this study concludes that exclusively and non-exclusively breastfeeding children in the study area are remarkably at higher risk of carcinogenic exposure compared to other regions.

### Aflatoxin M1 in the urine of breastfeeding children in Kibwezi West

The prevalence of aflatoxin M1 in the urine of breastfeeding children in this study was 100%. It was higher than 79% and 83% reported in Makueni and Nandi, respectively, for children below 5 years [23]. It was also higher than 17% in Ethiopia [46] for infants 1-2 years, 98.8% in Ogun Nigeria for children below 2 years [47], 47% in Colombia for infants [38], 53% in southern Ethiopia for children 6-23 months [48], and 43.5% in Bangladesh [49] for infants. The rates were also higher than the 4% and 12% rates reported for exclusive and non-exclusively breastfed children, respectively, in Nigeria [32]and the 22% rate reported for exclusively breastfed children in Iran [21]. The results reflect a high dietary intake of aflatoxin by lactating mothers in Makueni and mothers’ dietary role in influencing aflatoxin M1 in breast milk in the study area.

Based on concentration, the overall mean of aflatoxin M1 in the urine of breastfeeding children in this study (390 ng/L) was lower than 1182.9 and 857.3 ng/L for children below 30 months in Makueni and Nandi, respectively [23]. They were higher than 270ng/L (60-510 ng/L) in Nigeria [47], and 214 (0-2582 ng/L) [48] and 64 ng/L in Ethiopia [46]. The levels were also higher than 16 ng/L in Columbia [38], 55.6 ng/L in Bangladesh [49], and 39 ng/L reported in Sweden [50]. The mean for non-exclusive breastfed children in Nigeria (166 ng/L) [32] was lower than that reported in this study (420 ng/L). Likewise, exclusively breastfeeding children (23 ng/L) was lower than 350 ng/L reported in this study. Mean levels (96 ng/L) reported in Iran for exclusive breastfed [21] were also lower than those reported in this study. High levels of aflatoxin M1 in urine in this study were expected and confirmed that lactating mothers and breastfeeding children in Kibwezi are exposed to high levels of aflatoxin intake through maize-based foods. These results also reaffirm earlier sentiments of this study that aflatoxin occurrence is persistent in Makueni. However, significant variation of aflatoxin M1 noted between urine of non-exclusively breastfeeding children age groups could result from different complementary foods used before collecting urine samples.

### Factors associated with the occurrence of aflatoxin M1 in the breast milk of lactating mothers

In the study, total aflatoxin was the major predictor of aflatoxin M1 concentration levels in the breast milk of lactating mothers. It accounted for 71% of aflatoxin M1 levels in breast milk. Significant linearity was observed for mothers who exclusively and non-exclusively breastfeed their children. Estimates of aflatoxin B1 intake in the study were also a major influencer of aflatoxin M1 levels in breast milk. However, this was reported for exclusively lactating mothers as opposed to non-exclusively lactating mothers. This one-sided influence could not be explained since a nonsignificant difference in aflatoxin B1 intake was reported between exclusively and non-exclusively lactating mothers. However, the co-occurrence of aflatoxin B1 and M1 in breast milk and the complex metabolization process of aflatoxin in the body could not be ruled out. That notwithstanding, these results agree with studies by Kang’ethe et al. [23] among children below 5 years of age in Makueni, Kenya. Results in Nigeria were also consistent with the results of this study [51]. The results were also similar to those of Azarikia et al. [17] in Iran, Elaridi et al. [16] in Lebanon, and Mehta et al. [18] in India.

A predictive model containing mothers’ dietary diversity and aflatoxin weekly consumption score predicted about 32% of aflatoxin M1 intake among breastfeeding children in the study. This model emphasizes the importance of diverse diets in areas prone to aflatoxin contamination. It also points out that the type of food constituting a diverse diet is vital in reducing aflatoxin exposure among lactating mothers. Going by these results, maize-based foods are still considered the most significant dietary contributor of aflatoxin M1 in breastmilk of lactating mothers in the study area (maize *ugali* > maize porridge > maize sorghum porridge > *‘githeri’* > *‘muthokoi’*).

Earlier results of this study showed a direct role of household size, education level, and socioeconomic status on levels of aflatoxin intake among lactating mothers. On the contrary, in addition to the age of mothers and breastfeeding children in the study, a direct influence on aflatoxin M1 in breast milk was not observed. Consequently, no direct influence was observed on aflatoxin M1 intake among breastfeeding children. These observations are similar to those of Mehta et al. [18] and Elaridi et al. [16]. However, they differ from Karayağiz and Özdemir study [52], which reported a positive significant association.

Among variables of breastfeeding practices in the study, time for initiating breast milk, number of children per lactating mother, and time for introducing complementary foods to children did not directly influence children’s exposure to aflatoxin M1 through breast milk. This is worth noting since introducing children to complementary foods is expected to reduce suckling and, thus, breast milk intake. Also, it is impossible to statistically determine the correlation between breast milk intake and aflatoxin intake of M1. However, it is evident in this study that children who are most frequently breastfed will be exposed to higher levels of aflatoxin M1 in breast milk. These remarks point to the need for having an elaborate plan in the study area to reduce aflatoxin exposure among lactating mothers. This is because breast milk is the only food considered safe for children under six months [1].

Similarly, a significant association was not observed between maize source, handling, and storage practices with the occurrence of aflatoxin M1 in breast milk. However, this present study suggests the need to conduct robust research that links the food supply chain with the occurrence of aflatoxin in breast milk in the study area. This is because studies by Nabwire et al. [53] and Daniel et al. [54] have linked maize source, handling, and storage practices with aflatoxin contamination in Makueni and its surroundings.

### Factors associated with aflatoxin M1 in the urine of breastfeeding children

This study confirmed that total aflatoxin in foods influenced aflatoxin M1 in the urine of exclusively and non-exclusively breastfeeding children in the study area. It explained that about 15% of aflatoxin was in the urine of all children whose samples were analyzed. Also, a model containing total aflatoxin, age of children, and socioeconomic status was able to explain 70% of aflatoxin M1 in the urine of exclusively breastfeeding children. This study thus concludes that maize *ugali*, indirectly, is the most significant contributor of aflatoxin M1 in the urine of breastfeeding children in the study area. This is followed by maize porridge, maize sorghum porridge, ‘*githeri,’* and *‘muthokoi.’* Even though insignificant, a positive correlation between intake of total aflatoxin, aflatoxin B1, and aflatoxin M1 in breast milk with aflatoxin M1 in the urine of children is still supportive of Alegbe et al. [55] findings. The findings showed that mothers’ dietary intake patterns positively correlated with aflatoxin exposure. However, the absence of a direct link between aflatoxin B1 and aflatoxin M1 in urine in the study was due to the reasons stated by Ali et al. [49] and Boshe et al. [48] findings. They concluded that only a tiny percentage of aflatoxin B1 can be transferred to urine. Also, a significantly higher concentration level of aflatoxin M1 in the urine of non-exclusively breastfeeding children than that of exclusively breastfeeding children (*p* = 0.035) was noted. This difference pointed out that the use of complementary foods in the study area also influences aflatoxin M1 levels in the urine of breastfeeding children. This comparison is based on the findings of Ezekiel et al. [32] and Magoha et al. [20], who determined levels of aflatoxin among exclusively and non-exclusively breastfeeding children in Nigeria and Tanzania. They concluded that non-exclusively breastfeeding children are predisposed to higher levels of aflatoxin (breastmilk + complementary foods) than their counterparts who only consume breastmilk. Children’s age was not a significant predictor of aflatoxin M1 in the urine of exclusively breastfeeding children in the study. Despite this, a positive correlation intimated that an increase in age was associated with an increase in aflatoxin exposure. This was in parallel with the findings of this study that showed that non-exclusive breastfeeding rates increased with an increase in the age of children. This study, therefore, maintains that the age of children is an underlying factor in determining the extent of aflatoxin M1 exposure in the study area.

The negative correlation between socioeconomic status and aflatoxin M1 in the urine of exclusively breastfeeding children contradicted earlier findings of this study. The reports showed a positive correlation between socioeconomic status and aflatoxin B1 intake among exclusively lactating mothers. This contradiction reaffirms earlier sentiments of this study. It was suggested that socioeconomic status may not be a reliable pointer to aflatoxin exposure in an area where contamination is highly prevalent. Also, a direct influence of household size, age, and education level of lactating mothers was reported on aflatoxin intake in the study. On the contrary, this was not the case for aflatoxin M1 levels in the urine of breastfeeding children in the study. These findings were similar to those of Chen et al. [56].

Similarly, a direct influence of women’s dietary diversity, aflatoxin weekly consumption score, and the total number of daily meals on mothers’ aflatoxin intake were reported. Again, this was not the case for aflatoxin M1 in the urine of breastfeeding children in the study. These results were again similar to Chen et al.’s [56] among children in Tanzania. Again, this study could not show any influence of sourcing, handling, storing, and processing of maize on aflatoxin M1 in the urine of breastfeeding children. This is despite studies by Nabwire et al. [53] and Daniel et al.[54] linking various sources of maize and handling practices with aflatoxin contamination in the study area. A possible explanation for this was that this study did not determine the source of the analyzed food samples. Again, maize handling and storage practices in this study were only reported by mothers and not based on laboratory analysis results. However, there are no studies that have explored the relationship between maize handling and storage practices with the presence of aflatoxin in breast milk and the urine of breastfeeding children less than six months of age.

### Effect of aflatoxin exposure on weight-for-age of exclusively and non-exclusively breastfeeding children below 6 months in the study

The prevalence rate of underweight for exclusively and non-exclusively breastfeeding children was 13.3 and 11.6%, respectively. They were comparable to 13% and 18% reported in Tanzania among exclusive and non-exclusive breastfeeding children, respectively [20]. The overall prevalence rate (12.4%) for this study was almost similar to 10.2 and 11% reported for Makueni and Kenya, respectively [57], but slightly lower than the 14.6% reported by Kang’ethe et al. [23] also in Makueni. Despite comparable underweight rates, this present study did not show any direct influence of aflatoxin exposure on the weight-for-age outcome of breastfeeding children. This was contrary to the results of Kang’ethe et al. [23] and Wangia-Dixon et al. [24] who showed a negative association between aflatoxin exposure and weight-for-age among children aged below 5 years and 6 and 12 years, respectively, in Makueni. However, one striking difference between the studies mentioned earlier and this present study was the difference in children’s age. The studies mentioned above included older children, for whom this present study presumes exposure to more aflatoxin intake than breastfeeding children. However, Magoha et al. [58]with children aged six months and below, also reported a negative association between aflatoxin exposure and weight-for-age z-scores. Further analysis, however, revealed that the exclusive breastfeeding rate reported in this present study was higher than that of Magoha et al. [58]. Moreover, aflatoxin M1 levels in Magoha et al. [58] were higher than the ones reported in this study. From these results, it may be concluded that children who are not being breastfed or are non-exclusively breastfed are more exposed to aflatoxin intake than those who are exclusively breastfed. This is because most of the complementary foods being used in aflatoxin-prone areas have also been mentioned to be highly susceptible to aflatoxin contamination. These sentiments explain the reason why underweight prevalence rates (14.6%) reported by Kang’ethe et al. [23] among children below 5 years, 17% reported by Ayelign et al. [46] rate among infants in Ethiopia, and 17 and 21% reported by Chen et al. [56] among children aged 24 and 36 months, respectively, in Tanzania were slightly higher than the ones reported in this study.

Further analysis revealed that age was the only predictor of weight-for-age z-scores of breastfeeding children in this study. This sentiment agreed with Hoffmann et al. [59] who also concluded that the impact of aflatoxin exposure on growth parameters in children varies with age. However, other variables, including the age of lactating mothers, household size, education level, and socioeconomic status, did not directly impact the study’s weight-for-age z-score of breastfeeding children. Similarly, no direct influence of dietary consumption patterns and breastfeeding practices on weight-for-age z-scores was reported. However, while no direct effect of aflatoxin exposure on weight-for-age of exclusively and non-exclusively breastfeeding children was reported, this present study still emphasizes the need for exclusively breastfeeding children aged six months and below in the study area. This study further points out that the absence of a direct impact of aflatoxin exposure on weight-for-age does not rule out imminent adverse effects of chronic aflatoxin exposure in children’s later life. Studies including [19, 59, 60] have shown negative impacts of aflatoxin. It is, therefore, probable that breastfeeding children in this study area, due to high aflatoxin exposure, will be at risk of stunting, cancer, increased morbidities, and micronutrient deficiencies, among other adverse side effects. Because of this, this study recommends further follow-up and risk characterization. Clinical studies can help ascertain the impact of high aflatoxin exposure besides depending on weight-for-age z-scores in the study area.

## Conclusion

The high prevalence and concentration levels of aflatoxin in breast milk and urine samples indicate that children between 0 and 6 months in the study setting are highly exposed to aflatoxins through both breast milk and pre-lacteal feeds and suggests an under-evaluated risk in aflatoxin-prone regions. This present study did not show any direct influence of aflatoxin exposure on the weight-for-age outcome of breastfeeding children of 0-6 months, contrary to the results of studies conducted with older children.

### Recommendation

Lactating mothers should continue to follow WHO breastfeeding recommendations since this study shows that non-exclusively breastfeeding children in aflatoxin-prevalent regions face more risk of aflatoxin from pre-lacteal feeds susceptible to aflatoxin contamination. Despite the lack of immediate impact on aflatoxin exposure on weight-for-age breastfeeding children, we consider the duration below six months to be a short period to detect any significant changes in the weight-for-age parameters. As such, we do not rule out imminent adverse effects of chronic aflatoxin exposure in children’s later life. We recommend further follow-up and risk characterization studies using other indicators, including height-for-age z-scores, for more conclusive results.

## Data Availability

The data is available on figshare platform as a dataset (Aflatoxin in food, breast milk of lactating mothers, and urine of children below 6 months and associated data). DOI:10.6084/m9.figshare.28247918

https://doi.org/10.1111/mcn.13493

## Acknowledgments

We thank Kimani Gathombi (Bora Biotech Laboratory) and the Public Health Pharmacology and Toxicology Laboratory (University of Nairobi) for the laboratory analysis. We also acknowledge the support of Prof. Cheminingw’a Ndiema George (Turkana University College, Kenya) and the late Obimbo Lamuka (University of Nairobi) in conducting this study.

## Supporting information

S1 Table: Quantities of breastmilk consumed by breastfeeding children per age group in Kibwezi West

S2 Table: Weight-for-age z-scores of breastfeeding children by age group and breastfeeding status in Kibwezi West

S3 Table: Aflatoxin M1 intake in breastmilk by age group of breastfeeding children in Kibwezi West

S4 Table: Aflatoxin M1 intake in breast milk by breastfeeding status and age group of breastfeeding children in Kibwezi West

S5 Table: Margin of exposure of breastfeeding children to aflatoxin M1

S6 Table: Aflatoxin M1 in the urine of exclusively and non-exclusively breastfeeding children in Kibwezi West

